# Noninvasive Wound Swab Proteomics Enables Prognostic Stratification of Diabetic Foot Ulcer Healing: The HEAL-DFU Proof-of-Concept Study

**DOI:** 10.64898/2026.09.15.26363101

**Authors:** Marc Clos-Garcia, Siddhi Y. Jain, Cilia H. Naaman, Jana A. Raguz, Zeinab Schaefer, Gurjeet Kaur, Viktor R. Curovic, Klaus Kirketerp-Møller, Jonas A. Andersen, Susanne Engberg, Anne Rasmussen, Aristidis Veves, Peter Rossing, Tarunveer S. Ahluwalia

**Author notes:** **Corresponding author:**Tarunveer Singh Ahluwalia, PhD. Senior Researcher & Associate Professor, Steno Diabetes Center Copenhagen, Borgmester Ib Juuls Vej 83, DK-2730 Herlev. Equal contribution (Marc Clos-Garcia, Siddhi Y. Jain).

## Abstract

**Objective:** Diabetic foot ulcers (DFUs) affect up to 34% of people with diabetes and precede 80% of lower extremity amputations, yet no ulcer-level molecular biomarkers distinguish healing trajectories. We aimed to identify wound swab proteins associated with DFU healing time and characterize their prognostic potential.

**Research Design and Methods:** LC-MS/MS proteomics was performed on wound swabs from 112 people with diabetes presenting with new DFUs, followed for up to 6 months. Participants were divided into fast-healers (≤40), slow-healers (41-180 days) and chronic wounds (>180 days). Proteins associated with “time to healing” were identified using Firth-penalized Cox regression (FDR<0.2; HR<1 indicated slow healing) and used to construct a cross-validated Protein Healing Score (PHS). Sensitivity and causal mediation analyses examined clinical variable effects on protein abundance, with independent dataset validation.

**Results:** Participants had a mean (±SD) age of 67.5 (±12.3) years, and diabetes duration of 27.5 (±14.9) years. Twenty-seven proteins formed two clusters: a slow-healing cluster enriched in neutrophil granule and innate immune proteins (HR<1), and a fast-healing cluster enriched in proteostatic stress proteins (HR>1). Triglyceride levels differed between healing groups (p=0.001); seven slow-healing proteins showed triglyceride-mediated effects (21–28%), suggesting a dyslipidaemia–innate immunity axis. The PHS significantly stratified prognosis (adjusted HR=1.47, 95%CI 1.14–1.89; p=0.003), outperforming clinical variables alone (AUC-OVR 0.71 vs. 0.62). Transcriptomic validation confirmed directional concordance.

**Conclusions:** Wound swab proteomics at DFU onset identified a 27-protein healing signature linked to neutrophil–innate immunity and triglyceride associated dyslipidaemia, offering a potential non-invasive prognostic tool.

**Highlights:**

- Why did we undertake this study? No prior study has examined wound swab protein associations with DFU healing time as a continuous outcome, offering finer prognostic resolution than discrete healing categories.
- What is the specific question(s) we wanted to answer? Can wound swab proteins at DFU presentation predict healing trajectories, and do clinical factors mediate these associations?
- What did we find? Twenty-seven proteins formed two prognostic clusters: a slow-healing cluster enriched in neutrophil/innate immune proteins and a fast-healing cluster linked to proteostatic stress. A Protein Healing Score outperformed clinical variables in stratifying healing outcomes (AUC-OVR 0.71 vs. 0.62). Triglyceride-mediated and neuropathy-driven effects on wound proteins revealed mechanistic links to impaired healing.
- What are the implications of our findings? Wound swab proteomics offers a non-invasive prognostic tool at DFU onset, potentially enabling earlier targeted intervention.

---

Diabetic foot ulcers (DFUs) are a severe complication of diabetes characterized by impaired healing, persistent inflammation, and high risk of infection. Lifetime risk of incurring a DFU is estimated at 19-34%, with more than 50% of ulcers becoming infected (1,2) and ∼20% progressing to lower extremity amputations (LEA) (2–5). Non-healing DFUs carry a five-year mortality rate comparable to many malignancies and constitute the most resource-intensive diabetes complication globally (5).

DFUs were classified according to underlying etiology (predominantly neuropathic, ischaemic or neuroischaemic) (6–8): neuropathic ulcer (foot pulse, toe blood pressure >70mmHg, neuropathy); neuroischemic ulcer (foot pulse, toe blood pressure 40-70mmHg, neuropathy); and ischemic ulcer (foot pulse, toe blood pressure <40mmHg, no neuropathy). Peripheral neuropathy causes loss of protective sensation and insensitivity to repetitive trauma, while peripheral arterial disease impairs tissue perfusion and healing. Despite systematic neuropathy and vascular screening, no validated ulcer-level molecular biomarkers exist to stratify patients by healing trajectory at the point of care. Immune dysregulation, driven by hyperglycemia and its downstream effects on neutrophil and macrophage function, is increasingly recognized as a central mechanism in impaired DFU healing (9), thus suggesting a new target route for biomarker discovery. Healing of a diabetic foot ulcer is a dynamic and highly coordinated process involving hemostasis, inflammation, tissue proliferation, extracellular matrix remodeling and tissue maturation (9,10). An appropriate and timely immune response is essential for progression through these phases, with neutrophils and macrophages playing central roles in pathogen clearance, removal of damaged tissue, and initiation and resolution of inflammation (9,11–13). In diabetes, chronic hyperglycemia and its downstream metabolic effects can impair immune cell function and disrupt the transition from inflammation to tissue repair (9,14,15). Altered neutrophil and macrophage recruitment, activation, and function may contribute to persistent inflammation and impaired wound healing (11–13,16). These complex interactions between metabolic disturbances, immune dysfunction, vascular insufficiency, and neuropathy make DFU healing difficult to predict using clinical characteristics alone (1,9,17). They also provide a strong rationale for omics-based approaches that can capture molecular signatures of impaired healing and potentially identify biomarkers for early risk stratification and therapeutic targeting (18–22) .

Wound swab proteomics offers a non-invasive approach to capture the functional molecular landscape of the wound microenvironment. Swabs can be obtained as part of routine clinical wound assessment and yield quantifiable protein signatures without the need for biopsy. We hypothesized that proteome-wide profiling of DFU wound swabs collected at DFU diagnostics, combined with longitudinal follow-up of healing outcomes, would reveal mechanistic pathways of impaired healing and provide prognostic information beyond established clinical risk factors. Thus, the *HEAL-DFU* study (Healing Evaluation and Associated molecular signatures in Diabetic Foot Ulcers) investigated whether wound swab proteomics could identify mechanisms and prognostic biomarkers of healing.

## RESEARCH DESIGN AND METHODS

### Study Participants, Diagnosis, and Biobanking

Adult individuals with type 1 or type 2 diabetes presenting with newly diagnosed DFUs at the Steno Diabetes Center Copenhagen prospectively enrolled in the HEAL-DFU study in 2018. Diabetes diagnosis was made using the WHO criteria (23). A DFU was defined as a lesion of the skin on the foot of the person with diabetes (24). All information on DFUs was attained in accordance with the international classification of diseases, tenth revision (25).

Participants were followed for up to six months or until DFU healing, whichever occurred first. Data was collected at baseline and planned follow up visits. Information on clinical characteristics and DFU measures were extracted from the patient’s electronic health records. Time to DFU healing was the primary outcome investigated in the current study. Clinical healing groups were defined using clinically informed thresholds reflecting early or fast-healing (≤40 days), delayed or slow-healing (41–180 days), and chronic or non-healing (> 180 days) (26–28). Participants were followed for up to six months or until DFU healing, whichever occurred first. . Wound swab samples were collected at DFU diagnostics onset using standard cotton swabs (BD ESwab Collection and Transport System, Becton Dickinson & Co. USA) by rubbing the wound surface and stored at −80°C. The HEAL-DFU study (also referred to as *Micropredict*) was approved by the Regional Ethics Committee and all participants provided written informed consent (VEK H-1708891).

### Wound Swab Shotgun Proteomics

Whole lysates of participants ulcer swab samples were prepared using a guanidine HCL lysis buffer, as described in Jersie-Christensen *et al*(*29*). Protein concentration was measured on Nanodrop, and samples were digested with trypsin protease. Peptides (5µg) were subjected to high–resolution mass spectrometry LC/MS analysis. Raw data was quantified using MaxQuant v1.6.12.0, and LFQ values were used for data annotation and initial statistical analysis employing

Quality filtering using Perseus (version 2.0.10) was performed, retaining proteins with at least 30% non-missing values across the cohort. LFQ intensities were log2-transformed and z-score standardized for downstream modelling. Full preprocessing details are provided in the Supplementary Methods.

### Protein Association with *Time to Healing*

Continuous covariates and protein abundances were standardized (z-score). Missing clinical data on covariates were imputed by k-nearest neighbor (kNN) imputation (k=5).For each protein, a separate Firth-penalized Cox proportional hazards model was fitted, adjusted for age, sex, BMI, diabetes type, diabetes duration, and HbA1c measures at baseline. The modeled event was achievement of wound healing (event=1); participants who did not achieve healing were censored at 180 days (event=0, time=180). Because the event of interest is a favorable outcome (healing), hazard ratios are interpreted in the opposite direction to conventional survival models: hazard ratio (HR) >1 indicates an increased hazard of healing at any given time corresponding to a faster time to healing while HR<1 indicates a reduced hazard of healing, corresponding to a longer time to healing. *p*-values were adjusted using the Benjamini-Hochberg false discovery rate. Significance was defined as FDR < 0.2, appropriate for the discovery nature of this manuscript.

Functional enrichment was performed using *clusterProfiler* (R) against GO:Biological Process, with pathway clustering using the *aPEAR* network approach.

We defined significance threshold at FDR 0.2, appropriate for the discovery nature of this manuscript.

### Clinical Variables Association and Sensitivity Analyses

Spearman rank correlations were computed between significant proteins and clinical covariates, and among significant proteins themselves, to characterize co-expression structure. Based on the outcome of such analyses and available literature (14,30), sensitivity and mediation analyses were performed for the relevant variables (Supplementary Methods).

### Cross-Validated Protein Healing Score

To eliminate in-sample optimism, the Protein Healing Score (PHS) was computed by 5-fold stratified cross-validation using the differential proteins. In each fold, Firth-penalized Cox regression was refitted on training patients only, and z-score normalization parameters were estimated from training data and applied to held-out patients. PHS for each held-out patient was: PHS = Σ(β□ × z-score□), where β coefficients came from the training fold. Participants were assigned to three groups by k-means clustering (k=3) on the PHS distribution to mirror the original clinical grouping. Prognostic performance was assessed by Kaplan-Meier analysis (log-rank test), chi-square and Cramér’s V against clinical healing groups, and Cox regression adjusted for clinical covariates.es. Linear regression models were computed to determine the continuous effect of the PHS upon clinical days.

### Machine Learning Classification

Machine learning (ML) models TabPFN (transformer-based tabular foundation model suited to small clinical datasets) (31) and XGBoost were evaluated across five feature sets: (i) clinical variables only (baseline); (ii) 27 DEPs only; (iii) PHS only; (iv) clinical + DEPs; (v) clinical + PHS. Models were trained to distinguish each healing group (fast healing, slow healing and chronic non-healer ulcers) from the other groups (Supplementary Methods).

### External Validation

External proteomics validation was done by comparing our results to those presented in other DFU-based proteomics studies (18,19). Directional concordance between DEP associations and transcriptomic data was assessed in two independent datasets: GSE199939 (DFU vs. healthy skin; n=10 DFU, n=11 controls) (20) and GSE134431 (healer vs. non-healer DFU; n=13) (11), both analyzed by DESeq2 with Benjamini-Hochberg correction (Supplementary Methods).

## RESULTS

### Study Cohort and Clinical Characteristics

The study included 112 participants with DFU with a mean (SD) age of 67.5 (±12.3) years; 80.4% were male and 62.5% had type 2 diabetes (Table 1). Three clinical groups were defined according to their DFU healing time: fast-healers, including people that healed under 40 days (n=48); slow-healers, including individuals healing within 41 to 180 days (n=46); and chronic wounded, not healing after 180 days (n=23). The mean time to heal for the fast-healers was of 22.5 (±10.9) days and 88.2 (±37.1) days for the slow-healers. Median diabetes duration was 24 (±15.17) years and 83.9% had peripheral neuropathy. The three healing groups were comparable across most clinical variables (Table 1). Triglyceride levels differed significantly across groups, increasing progressively with longer time to healing (fast-healing: 1.7±0.9; slow-healing: 2.0±1.0; chronic: 2.9±1.9 mmol/L; P=0.001). Wounds were identified at ankle (malleolar ulcer, n=2), dorsal surface (n=11), foot bed (plantar ulcer, n=24), heel (n=8) and toe (n=67), being the wound placement significantly different across healing categories (χ² *p*=0.003). Wound area and volume were significantly larger in chronic wounds (P=0.011 and P=0.027 vs. fast healing, respectively).

**Table 1.** Clinical characteristics of participants compared between the three healing groups. Continuous variables represented as Mean (Variance) and were tested using F-test (ANOVA) and categorical variables are represented as Total number (%) and were tested using Chi-square test. *P<0.05. FH, fast-healing; SH, slow-healing; C, chronic; TBI, toe-brachial index. For the days-to-heal variable, NA indicates no healing within 180 days of follow-up (censored event).

| Variable | Fast-healing<br>(n=47) | Slow-healing<br>(n=42) | Chronic<br>(n=23) | Total (n=112) | P value |
| --- | --- | --- | --- | --- | --- |
| Days to heal | 22.5 (10.9) | 88.2 (37.1) | NA | 53.5 (42.3) | <0.001* |
| Age, years | 67.1 (11.4) | 66.2 (12.8) | 70.6 (13.1) | 67.5 (12.3) | 0.37 |
| Sex, male, n (%) | 36 (76.6) | 36 (85.7) | 18 (78.3) | 90 (80.4) | 0.54 |
| Type 1 diabetes, n (%) | 23 (48.9) | 12 (31.0) | 6 (26.1) | 42 (37.5) | 0.10 |
| Type 2 diabetes, n (%) | 24 (51.1) | 29 (69.0) | 17 (73.9) | 70 (62.5) |  |
| Diabetes duration, years | 29.9 (16.2) | 25.3 (13.5) | 26.4 (14.3) | 27.5 (14.9) | 0.37 |
| BMI, kg/m<sup>2</sup> | 30.5 (4.8) | 29.1 (5.9) | 30.1 (6.3) | 29.9 (5.5) | 0.59 |
| HbA1c, mmol/mol | 64.9 (14.0) | 65.3 (17.4) | 63.9 (14.2) | 64.8 (15.3) | 0.94 |
| Insulin use, n (%) | 38 (80.9) | 36 (85.7) | 22 (95.7) | 96 (85.7) | 0.25 |
| Peripheral neuropathy, n (%) | 41 (87.2) | 37 (88.1) | 16 (69.6) | 94 (83.9) | 0.11 |
| Triglycerides, mmol/L | 1.7 (0.9) | 2.0 (1.0) | 2.9 (1.9) | 2.0 (1.3) | 0.001* |
| TBI | 0.4 (0.2) | 0.5 (0.3) | 0.5 (0.2) | 0.5 (0.2) | 0.62 |

### Proteomics Identifies Two Biologically Coherent Healing Clusters

LC-MS/MS identified 1,092 proteins across 112 samples (Supplementary Table S1), of which 256 were retained after quality filtering (Supplementary Table S2). Firth-penalized Cox regression identified 27 proteins significantly associated with time to healing at an FDR threshold of <0.2 (Figure 1A, Supplementary Table S3), including PSMA5 (HR 1.45), CORO1A (HR 0.69), S100A8 (HR 0.68), and PGD (HR 0.65), S100A12, GSN, and BPI.

**Figure 1.**
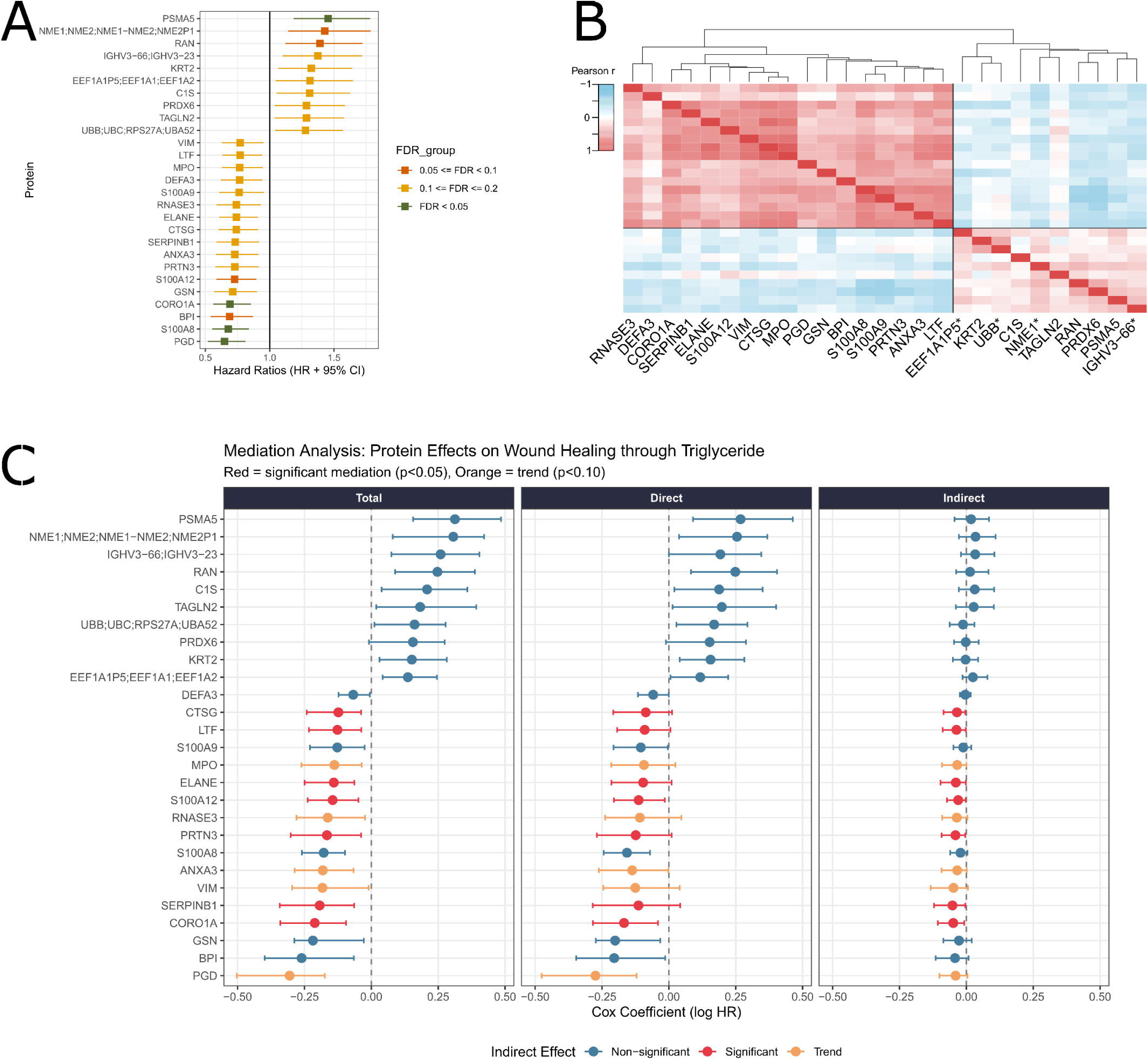
Proteins associated with DFU Time to healing, functional context and triglycerides mediation. (A) Forest plot of hazard ratios (HR) from Firth-penalized Cox regression models, estimated separately for each of the 27 proteins significantly associated with DFU time to healing (FDR ≤ 0.20). Each square represents the HR point estimate and horizontal bars represent the 95% confidence interval. Proteins with HR<1 are associated with slower healing, while proteins with HR>1 are associated with faster healing. Square color indicates the FDR threshold: dark green, FDR < 0.05; orange, 0.05 ≤ FDR < 0.10; yellow, 0.10 ≤ FDR ≤ 0.20. Proteins are ordered by descending HR. (B) Pairwise Spearman correlation matrix for the 27 differentially expressed proteins, visualized as a hierarchical clustering heatmap. Cell color encodes the correlation coefficient from −1 (blue) to +1 (red), as shown in the color key. Both rows and columns were clustered using complete linkage. (C) Forest plot showing total, direct, and indirect Cox regression log-hazard-ratio (log HR) coefficients for the 27 proteins significantly associated with DFU time to healing (FDR < 0.20). The Total effect represents the unadjusted protein-healing association; the Direct effect represents the protein-healing association independent of triglycerides; the Indirect effect represents the component of the association transmitted through triglyceride levels. All coefficients are expressed as log HR, where negative values indicate association with slower healing (HR < 1) and positive values with faster healing (HR > 1). Error bars represent 95% confidence intervals estimated from 1,000 bootstrap resamples. Proteins are ranked by their total effect estimate. Dot color encodes the statistical significance of the indirect (mediated) effect: red indicates significant mediation (p < 0.05), orange indicates a trend towards mediation (p < 0.10), and blue indicates no significant mediation. Proteins in the upper portion of the plot (positive log HR) belong to the fast-healing cluster (Cluster 1); proteins in the lower portion (negative log HR) belong to the slow-healing innate immune cluster (Cluster 2). Triglyceride levels were included as the sole candidate mediator. All models were adjusted for age, sex, BMI, and diabetes type. *EEF1A195 = EEF1A195;EEF1A1;EEF1A2, UBB = UBB;UBC;RPS27A;UBA52, NME1 = NME1;NME2;NME1-NME2;NME2P1, IGHV3-66 = IGHV3-66;IGHV3-23

Hierarchical clustering of pairwise protein-protein correlations revealed two clusters with strongly negative inter-cluster correlations (Figure 1B). Slow-healing cluster (**SH_C_**) (HR<1, n=17 proteins) comprised neutrophil granule proteins (ELANE, CTSG, PRTN3, MPO, RNASE3, DEFA3) and innate immune effectors (S100A8, S100A9, S100A12, LTF, BPI, ANXA3, SERPINB1, CORO1A, VIM, GSN, PGD), all of them negatively associated with healing. Faster-healing cluster (**FH_C_**) (HR>1, n=10 proteins) comprised markers of proteostatic stress and cellular damage: PSMA5 (proteasome subunit), UBB/UBC (ubiquitin), EEF1A1 (translation factor), PRDX6 (oxidative stress), NME1/NME2, RAN, KRT2, TAGLN2, C1S, and IGHV3-66, all of them positively associated with healing. GO:BP enrichment (130 significant pathways, Supplementary Table 4) was dominated by antimicrobial humoral response (−log_10_HMP=11.31) and neutrophil chemotaxis (−log_10_HMP=4.28) (Supplementary Figure 1-2, Supplementary Table S5) for SH_C_, and by UTP biosynthesis and translational elongation for FH_C_, consistent with the slow-healing cluster reflecting active innate immune effector function and the fast-healing cluster reflecting anabolic protein synthesis and turnover.

### Sensitivity analyses

We first tested how the 27 DEPs were associated with clinical data variables (Supplementary Table S6) to determine which sensitivity analyses were needed. Both triglyceride and neuropathic wound etiology were found to be relevant. A neuropathy sensitivity analysis (Supplementary Figure 3), adding wound etiology as a covariate to the Cox model, resulted in loss of significance for seven SH_C_ proteins (ANXA3, PRTN3, SERPINB1, MPO, VIM, S100A9, PRDX6), indicating that neuropathy etiology partially drives the SH_C_ associations in patients with intermediate healing trajectories.

Causal mediation analysis identified significant indirect effects through triglycerides for seven SH_C_ proteins (all P<0.05, Figure 1C, Supplementary Table S7) with proportions mediated ranging from 21.8% (S100A12) to 28.2% (LTF): CORO1A (23.0%; P=0.010), S100A12 (21.8%; P=0.024), ELANE (25.7%; P=0.034), CTSG (26.1%; P=0.016), PRTN3 (26.0%; P=0.022), LTF (28.2%; P=0.024), and SERPINB1 (26.9%; P=0.030). For all seven proteins, direct effects became non-significant after triglyceride adjustment, indicating full mediation. No FH_C_ protein showed triglyceride mediation. These findings were independently supported by significant positive correlations between triglyceride levels and CORO1A, S100A8, S100A9, and PRTN3 in the clinical dataset.

### Cross-Validated Protein Healing Score

The cross-validated PHS achieved statistically significant Kaplan-Meier stratification across three PHS-derived patient groups (log-rank P<0.0001; Figure 2A), where the median healing time between fast-healers and chronic wounded was 47 days. Agreement with clinically defined healing groups was moderate but significant (chi-square P=0.035; Cramér’s V=0.21; Figure 2B), with the PHS most reliably identifying fast-healing patients (68.2% correctly assigned in the high-PHS group) and least effective for chronic wounds (27.9% in the low-PHS group). A 3-fold difference was observed between the median time-to-heal between the highest PHS tertile (25 days to heal), corresponding to the fast-healer individuals, and the lowest PHS tertile (72 days to heal), the chronic wound participants. Linear regression showed that 1 SD increment on the PHS resulted in a reduction of 9.5 days (95% CI 0.8–18.2, *p*=0.033, R^2^=0.048) of healing time (Figure 2C). The moderate Cramér’s V indicates that the proteomic signature captures biological information partially independent of clinical classification, providing potential additive prognostic value.

**Figure 2.**
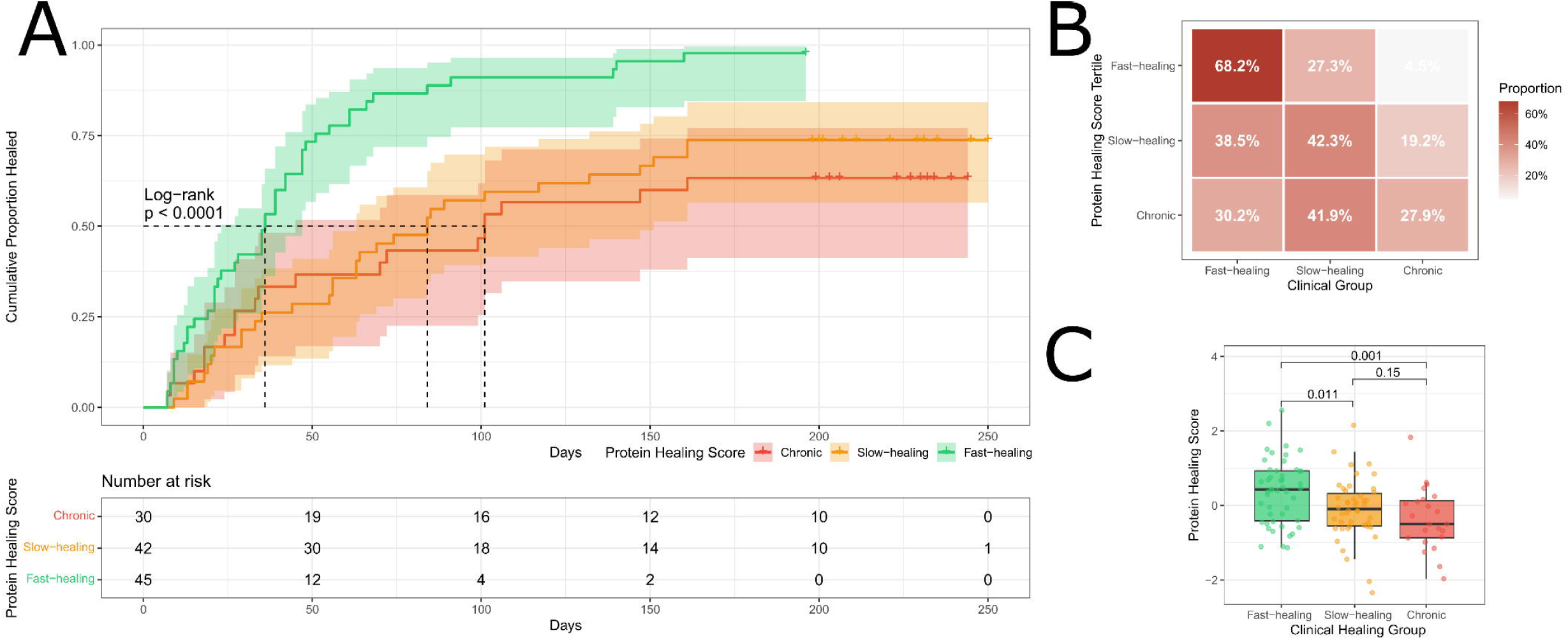
Protein Healing Score stratification of DFU healing trajectories. (A) Cumulative proportion of healed individuals for the three PHS-derived patient groups: Fast-healing (green), Slow-healing (orange), and Chronic (red). Group membership was assigned by k-means clustering (k = 3) applied to the continuous cross-validated PHS distribution. Shaded areas represent 95% confidence intervals. The dashed vertical lines indicate the median survival time for the Fast-healing group. Group separation was assessed by the log-rank test (p < 0.0001). The number-at-risk table below the plot shows patient counts at each time point for each PHS group. (B) Cross-tabulation of PHS-derived groups (rows) against clinically defined healing categories (columns), showing the row-wise proportion of patients assigned to each clinical group. Cell color encodes the proportion, from white (low) to dark red (high). Agreement between the two classification systems was assessed by chi-square test of independence (p = 0.0355) and Cramér’s V as an effect-size measure (V = 0.21), indicating modest but statistically significant concordance. (C) Distribution of continuous PHS values across the three clinically defined healing groups, shown as box plots with individual data points. The central line represents the median, box limits represent the interquartile range, and whiskers extend to 1.5× IQR. Pairwise differences between groups were tested using Wilcoxon rank-sum tests; p-values are shown for each comparison (Fast-healing vs Slow-healing: p = 0.011; Slow-healing vs Chronic: p = 0.001; Fast-healing vs Chronic: p = 0.15).

### Machine Learning Classification

Protein features outperformed clinical variables alone in both TabPFN and XGBoost classifiers (AUC-OVR 0.71 vs. 0.62; Figure 3, Supplementary Table S8). Adding clinical variables to the protein feature set offered no performance improvement.

**Figure 3.**
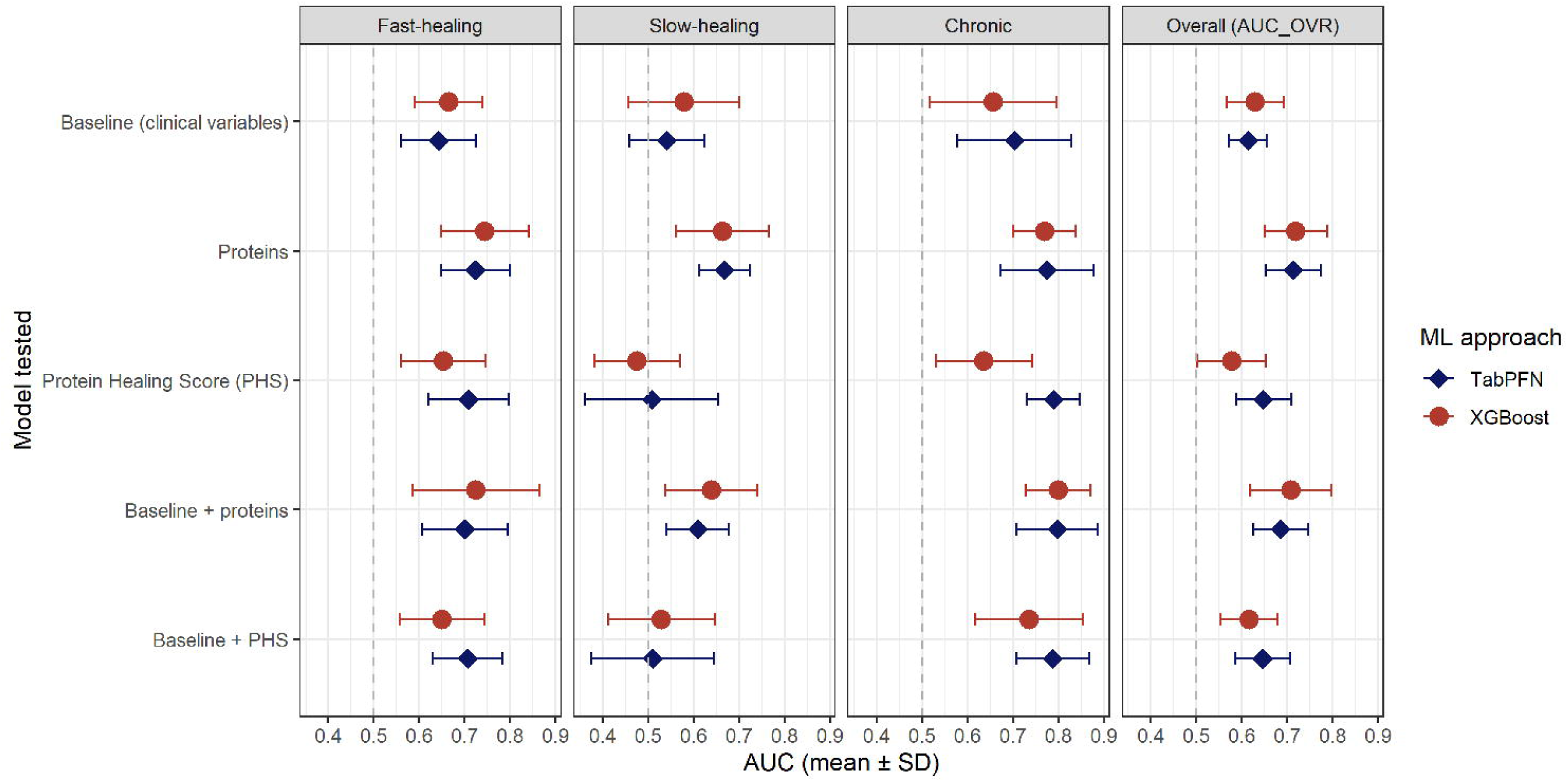
Machine learning classification performance across feature sets and healing groups. Forest plot of AUC (mean ± SD) for multiclass classification of DFU healing trajectory using two machine learning algorithms: TabPFN (dark blue diamonds) and XGBoost (red circles) across five feature sets. Each panel shows per-class AUC from a one-vs-rest (OVR) strategy for each of the three healing groups (Fast-healing, Slow-healing, Chronic); the rightmost panel shows the macro-averaged overall AUC-OVR summarizing multiclass discrimination across all three groups simultaneously. The five feature sets compared are: clinical variables alone (Baseline), wound swab proteins alone (Proteins), the Protein Healing Score alone (PHS), clinical variables combined with proteins (Baseline + proteins), and clinical variables combined with the PHS (Baseline + PHS). Error bars represent ± 1 standard deviation across 5-fold cross-validation repeats. The vertical dashed reference line at AUC = 0.5 indicates chance-level discrimination.

### External Validation

We identified 15 out of the 27 DEPs in the protein-protein interaction network provided in Wang *et al* results (19), even do Wang *et al* performed proteomics in a different context (healthy control *vs* DFU) and using a different technology. Specifically, they reported neutrophil degranulation and neutrophil activation as the dominant biological processes in the DFU wound proteome, overlapping with our slow-heal cluster proteins (S100A8/9/12, MPO, CTSG, PRTN3, BPI, CORO1A, VIM, LTF, DEFA3, SERPINB1, TAGLN2; Supplementary Table 3 and 11). Seemingly Soto *et al* (18) identified a small overlap with our results, particularly those associated with keratinocyte activation markers and extracellular vesicle-associated stress (MPO, PRTN3, CTSG, KRT2). Validation in transcriptomics datasets (11,20) and single-cell dataset (12) validated only a limited subset of proteins (Supplementary Results), coherently with the different omics approaches followed and sampled tissues.

## DISCUSSION

DFU wound healing is a complex biological process disrupted by several factors as hyperglycemia, chronic inflammation and neuropathy (10). Here, we present a multi-angle proteomics characterization of DFU swabs, identifying a 27-protein signature capable of stratifying people with DFU into biologically distinct healing trajectories and uncover a neutrophil-dyslipidemia axis as a central wound-healing mechanism. Furthermore, the PHS was found to be related to healing time, with 1SD increment reflecting a 9.5 days reduction in healing time. Notably, our results are partially replicated in both proteomics and transcriptomics external results.

The slow-healing cluster (SH_C_, 17 proteins) was characterized by neutrophil granule proteins and markers of the innate immune response at wound site. Higher wound abundance of neutrophil granule proteins (ELANE, CTSG, PRTN3, MPO) and calprotectin complex members (S100A8/A9/A12) in DFU with slower healing is consistent with a sustained or poorly resolving neutrophil response rather than the transient inflammatory response accompanying uncomplicated repair. Continued release of serine proteases and myeloperoxidase may contribute to extracellular matrix degradation and oxidative stress, providing a potential mechanism linking a high neutrophil protein burden at DFU diagnosis with prolonged healing (32,33). Other factors including persistent tissue damage related to inadequate off-loading or bacterial activity, may also contribute to sustained neutrophil activation; however, these mechanisms cannot be directly evaluated in the present study.

The biological identity of the SHC was supported across analytical approaches. Functional enrichment identified antimicrobial humoral response and neutrophil chemotaxis, while machine learning (ML) models consistently selected innate immune proteins among the most predictive features. In contrast, cell deconvolution identified relatively greater NK-cell and dendritic-cell signatures in DFU with faster healing. These findings are consistent with previous studies implicating neutrophil and macrophage dysfunction in impaired DFU healing (13,16).

Our findings also highlight the importance of biological compartment when integrating proteomic and transcriptomic data. Although S100A8/S100A9 have been identified as immune hub genes in DFU pathogenesis (21), scRNA-seq has reported enrichment of S100A8/S100A9-expressing M1 macrophages in healing DFU tissue (12), consistent with our pseudobulk transcriptomic findings for S100A8, S100A9, and PGD. This apparent discrepancy may reflect the different biological compartments captured by the two approaches: wound swabs measure extracellular proteins released into wound exudate, including products of neutrophil degranulation and cell death, whereas tissue transcriptomics primarily reflects gene expression in viable cells. Thus, high extracellular abundance of neutrophil-derived proteins and relatively high transcriptional activity in viable immune cells are not necessarily contradictory and may represent different stages or aspects of the inflammatory response. PGD emerged as one of the most consistently discriminative ML features. Its association with neutrophil metabolic activity is biologically plausible, although its precise role in DFU healing remains to be established. Paired assessment of wound proteins and tissue transcriptomes from the same DFUs, together with measures of bacterial activity and off-loading, will be required to further disentangle these mechanisms.

Despite sharing calprotectin scaffold (34), S100A12 showed significant triglyceride mediated effects (21.8%), whereas S100A8 and S100A9 did not suggesting that structurally related neutrophil proteins may be differentially influenced by metabolic factors. Similarly, the ELANE/SERPINB1 pair showed comparable triglyceride mediated effects (∼25-27%), supporting metabolic flexibility of neutrophil elastase regulatory axis. Overall, causal mediation analysis identified a dyslipidemia–innate immunity axis accounting for 21–28% of the protective protein effects on healing time, consistent with clinical correlations and high ranking of triglycerides in ML models. These findings suggest that triglyceride-associated modulation of neutrophil activity may link systemic metabolic dysfunction with the local wound environment, although causality cannot be established from the present data.

The fast-healing cluster was enriched in proteins involved in proteostatic, translation neucleotide metabolism and redox regulation. Although these pathways can be activated in response to cellular stress (15,35), their higher abundance in faster-healing DFUs suggests that this signature may instead reflect preserved cellular activity and tissue repair capacity. The enrichment of translation and UTP biosynthesis pathways, together with KRT2 and TAGLN2, is compatible with viable epithelial and contractile cellular compartments involved in tissue repair. However, the biological meaning of this proteomic pattern remains uncertain. Wound swab proteomics captures extracellular proteins present in wound exudate, whereas transcriptomic analyses of tissue primarily reflect gene expression in viable cells; therefore, differences between the two datasets may partly reflect compartmental differences. Our study cannot determine whether the detected proteins are actively secreted, released during physiological turnover, or associated with cell death. The fast-healing cluster was not materially affected by triglyceride adjustment, further suggesting that it represents a pathway distinct from the triglyceride-associated neutrophil signature. Previous work has shown that baseline inflammatory proteins in the wound environment are associated with subsequent DFU non-healing (36), supporting the broader relevance of wound proteomics for prognostic stratification.

Neuropathic etiology independently modulated the neutrophil signature, with seven proteins from the slow-healing cluster losing significance after adjustment for wound etiology. Neuropathic etiology was positively associated with abundance of these proteins, suggesting that part of the neutrophil signature associated with slower healing may be particularly prominent in neuropathic DFUs. This is consistent with the recognized contribution of peripheral and autonomic neuropathy to impaired wound repair through altered immune and neuropeptide-mediated signaling (17,22). Together with the triglyceride-associated effects described above, these findings suggest that metabolic and neurological factors may influence the same neutrophil-associated wound signature through distinct pathways. However, the present study cannot establish the underlying mechanisms. Cell deconvolution analyses provided further insight into the immune landscape associated with healing DFUs with slower healing showed relatively greater eosinophil-associated signatures and lower NK-cell and dendritic-cell signatures compared with faster-healing DFUs. Although these findings are exploratory, they are consistent with previous reports linking eosinophil-associated inflammation with chronic or non-healing wounds and suggesting roles for NK cells in immune surveillance and wound resolution (37–41). Together, these findings support the presence of distinct immune profiles across the healing trajectory, while emphasizing the need for cellular and functional validation.

The cross-validated PHS retained independent prognostic value after clinical covariate adjustment (HR=1.47; P=0.003), indicating that wound proteome provides information beyond established clinical risk factors. More importantly, it was found to be directly related to healing time, with increases in PHS value associated to healing time reductions. Protein features also outperformed clinical variables in machine learning classification, with no additional benefit from combining the two, suggesting that the 27-protein signature captures biological information not reflected by age, sex, BMI, diabetes type or duration, HbA1c, or triglycerides. The moderate association between PHS-derived and clinical groups (Cramér’s V=0.21) further supports partial independence between the proteomic and clinical information.

## Strengths and Limitations

A major strength was the prospective longitudinal design of a well-characterized cohort of people with newly diagnosed DFUs, enabling wound swab collection at DFU diagnosis and subsequent assessment of healing trajectories. Standardized biobanking and triplicate LC-MS/MS measurements further supported analytical robustness. The use of non-invasive wound swabs also provides a clinically scalable sampling strategy compatible with routine wound assessment.

Several limitations should be considered. The absence of an external proteomic cohort with longitudinal DFU healing data limited independent validation of the identified markers and PHS. The sample size (n=112) also resulted in uncertainty around machine-learning performance estimates. Defining chronic wounds as those not healed within 180 days may have introduced outcome misclassification and selection bias. Mediation analyses relied on assumptions of no unmeasured confounding, and the observational design precludes causal inference. Finally, comparisons with transcriptomic datasets should be interpreted cautiously because wound swab proteomics capture extracellular and exudate-associated proteins, whereas tissue transcriptomics reflects gene expression in viable cells. Overall, the consistency of findings across survival modelling, mediation analysis, machine learning, cellular deconvolution, and transcriptomic validation supports the biological relevance of the identified signatures. These findings demonstrate the potential of non-invasive wound swab proteomics to provide biologically informed prognostic stratification in DFU and warrant external validation in larger, multicentre cohorts. The findings of the current study highlight the potential of non-invasive wound swab proteomics to support biologically informed risk stratification and precision medicine approaches in diabetic foot care.

## Supporting information

Supplementary Results

## Data Availability

The data sets used in this study are available from the corresponding author after acquiring required permissions from the relevant regulatory authorities.
All required code for this manuscript replication is accessible at https://github.com/SDCC-ClinicalOmics/DFU_recovery_PX

https://github.com/SDCC-ClinicalOmics/DFU_recovery_PX

## Acknowledgments

We thank the participants in this study. We also thank Prof. Louise Dalgaard for facilitating the use of Proteomics core Lab. facility and Marija Petkovic for assisting in sample preparation at the Roskilde University for this project. We acknowledge Dr. Rosa Jersie-Christensen for her contributions to the study.

Tarunveer S. Ahluwalia is the guarantor of this work and, as such had full access to all data in the study and takes responsibility for the integrity of the data and the accuracy of the data analysis.

AI tools (Claude tools) were used to assist in code writing and manuscript text editing and formatting.

## Author Contributions

Conceptualization: TSA

Methodology: MC-G, SYJ, TSA

Investigation: MC-G, SYJ, JAR, RRJ-C, SE, AR, TSA

Visualization: MC-G

Funding acquisition: PR, TSA

Project administration: AR, TSA

Supervision: TSA

Writing – original draft: MC-G, SYJ, TSA

Writing – review & editing: MC-G, SYJ, CHN, ZS, GK, VRC, KK-M, JA, AV, PR, AR, TSA

All authors read and approved the final version of the manuscript.

## Data and code availability

The data sets used in this study are available from the corresponding author after acquiring required permissions from the relevant regulatory authorities.

All required code for this manuscript replication is accessible at https://github.com/SDCC-ClinicalOmics/DFU_recovery_PX

## SUPPLEMENTARY METHODS

The following details supplement the Research Design and Methods section of the main manuscript.

### Data Preprocessing

The LC-MS/MS analysis was performed three times per sample and values averaged across runs. Perseus (version 2.0.10) was used for quality filtering; only proteins with ≥30% non-missing values across all 112 samples were retained (n=256 proteins). Missing protein values were imputed from a normal distribution (width=0.3, shift=1.8 of the log-normal distribution) following Perseus defaults. LFQ intensities were log2-transformed by MaxQuant and z-score standardized prior to modelling so that hazard ratios reflect per-standard-deviation changes in abundance.

### Clinical Data Imputation

Missing clinical covariate values were imputed using k-nearest neighbour (kNN) imputation with k=5 using the VIM R package. Continuous variables (age, BMI, HbA1c, diabetes duration, log-triglycerides) were z-score standardised; categorical variables (sex, diabetes type) were dummy-encoded.

### Proteins association with healing time

A Cox proportional hazard model was fitted individually for each protein with Firth’s penalized likelihood correction with *coxphf* R package to address potential separation and convergence issues arising from the high-dimensional proteomic data. Each model was adjusted for age, sex, BMI, diabetes type, diabetes duration, and HbA1c. A neuropathy type (coded as neuropathic, ischemic-neuropathic, or ischemic) sensitivity analysis was performed, including neuropathy as a covariate where specified. Coefficients, 95% confidence intervals, and p-values were extracted for the protein term in each model. P-values across all protein-wise models were adjusted for the false discovery rate (FDR) using the Benjamini–Hochberg procedure. Due to the sample size and the discovery nature of the study design, proteins were considered statistically significant at FDR < 0.2.

### GO:BP Enrichment Analysis

Gene Ontology Biological Process (GO:BP) over-representation analysis was performed using clusterProfiler (R package) with the human annotation database (*org.Hs.eg.db*), applying Benjamini-Hochberg correction (α=0.05). Enriched terms were clustered using the *aPEAR* enrichment network approach based on pathway gene set overlap. Pathway-level directionality scores were computed based on the sign of constituent protein Cox HRs. P-values were summarized within clusters using the *harmonicmeanp* R package (harmonized mean p-value, HMP). Fold enrichment was computed as the ratio of observed to expected gene set overlap.

### Cell Type Deconvolution

To contextualize protein findings within the wound microenvironment, two complementary approaches were applied. (1) Single-sample Gene Set Enrichment Analysis (*ssGSEA*; *GSVA* R package) using DFU-specific scRNA-seq marker gene sets (12). All differentially expressed proteins (DEPs) were excluded from marker sets to ensure analytical independence from the survival analysis. Cell types with fewer than three detectable markers were excluded, retaining five cell types. (2) BayesDeBulk deconvolution (*proteoDeconv* R package) using a blood immune cell proteome reference (42). Estimated fractions were normalized to relative proportions within each sample.

### Single-sample GSEA Cell Type Scoring

Single-sample GSEA (ssGSEA) was performed using the *GSVA* R package (gsvaParam function). Cell type marker gene sets were derived from single-cell RNA sequencing of DFU wound tissue published by Theocharidis *et al*. (12), comprising the top 10 marker genes per cell type across 21 annotated populations. To ensure analytical independence from the Cox survival analysis, all 27 DEPs were excluded from the marker sets prior to scoring. Cell types with fewer than 3 detectable marker proteins in the proteomics dataset were excluded, retaining 5 cell types. Group differences were assessed by Kruskal-Wallis test with pairwise Wilcoxon rank-sum tests and Benjamini-Hochberg correction.

### BayesDeBulk Immune Cell Deconvolution

Reference-based deconvolution was performed using the BayesDeBulk algorithm (*proteoDeconv* R package). A proteomics-specific reference matrix was constructed from publicly available iBAQ intensity data from 28 primary human hematopoietic cell populations (Rieckmann et al., Nature Immunology, 2017). Cell subtypes were collapsed into 7 functional groups: neutrophils, monocytes, CD4+ T cells, CD8+ T cells, NK cells, eosinophils, and dendritic cells. Proteomics intensities were reverse log2-transformed and normalized to a TPM-like scale prior to deconvolution. Estimated fractions were converted to relative proportions within each sample (summing to 100%). Cell types with mean relative fraction <1% across samples were excluded. Statistical testing as for ssGSEA.

### Protein association to clinical data and mediation analyses

To assess whether the association between plasma proteins and wound healing time was mediated through circulating triglyceride levels, a causal mediation framework was applied to proteins passing the FDR ≤ 0.20 threshold. For each protein, three models were estimated:

- Total effect — a Cox proportional hazards model regressing the survival outcome on the protein, adjusting for age, sex, BMI, Hb1Ac, diabetes duration and diabetes type, with no mediator included.
- Alpha path (exposure → mediator) — a linear regression model with log-transformed triglycerides as the outcome and the protein as the predictor, adjusted for age, sex, BMI, Hb1Ac, diabetes duration and diabetes type.
- Direct effect and beta path — a Cox model including both the protein and log-triglycerides simultaneously, adjusted for age, sex, BMI, Hb1Ac, diabetes duration and diabetes type, yielding the direct effect of the protein on survival independent of triglycerides, and the beta path (triglyceride → survival).

The indirect effect (mediated through triglycerides) was estimated as the product of the alpha and beta path coefficients (α × β). Confidence intervals and two-sided p-values for the indirect effect were derived from 1,000 non-parametric bootstrap iterations (sampling with replacement). Bootstrap iterations with non-convergent models were excluded. The proportion mediated was calculated as the mean indirect effect divided by the mean total effect across bootstrap samples. Proteins with an indirect effect p < 0.05 were considered to have significant triglyceride-mediated effects; p < 0.10 was used as a trend threshold. All mediation models used the kNN-imputed clinical dataset with standardized covariates (as described above).

### Protein healing score computation and performance evaluation

A composite Protein Healing Score (PHS) was derived from the proteins passing the FDR ≤ 0.20 significance threshold in the Firth-penalized Cox models. Each significant protein abundance was first z-score standardized. To avoid optimistic performance estimates from in-sample evaluation, the PHS was computed using a cross-validated procedure. Patients were randomly partitioned into five stratified folds, preserving the distribution of the three clinical healing groups across folds. For each fold, Firth-penalized Cox regression was refitted exclusively on the training patients, and z-score normalisation parameters (mean and standard deviation) were estimated from training data only. The PHS for each held-out test patient was then computed as a weighted linear combination of the standardised protein values, where the weights were the Cox regression log-hazard-ratio coefficients (β) from the protein-wise models:

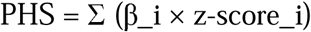

Patients were assigned to one of three score-based groups (chronic, slow-healing, fast-healing) using k-means clustering (k = 3) on the continuous PHS distribution (*classInt* package, R). Kaplan–Meier survival curves were estimated for each PHS group and compared with the log-rank test. Agreement between PHS-derived groups and the original clinically defined healing categories was assessed using a chi-square test of independence and Cramér’s V as an effect-size measure. Between-group PHS differences were tested with pairwise Wilcoxon rank-sum tests.

### Machine Learning Details

To evaluate the predictive value of the protein panel and the derived Protein Healing Score (PHS) for classifying wound healing outcome, two supervised machine learning models were benchmarked across five feature sets: (1) Baseline: seven clinical variables only (sex, age, BMI, diabetes type, diabetes duration, HbA1c, triglycerides); (2) Proteins: the panel of significant proteins (FDR ≤ 0.20) only; (3) ProteinScore: the PHS as a single continuous predictor; (4) Baseline + Proteins: clinical variables combined with the protein panel; (5) Baseline + PHS: clinical variables combined with the PHS.

The two models evaluated were: TabPFN (a transformer-based in-context learning classifier/regressor) and XGBoost (200 estimators). For clinical variables, remaining missing values were median imputed prior to modelling. Proteins with more than 50% missing values across samples were excluded; remaining missing protein values were median-imputed.

Classification performance (three healing groups: fast-healing, slow-healing, chronic) was assessed using stratified 5-fold cross-validation. Metrics reported were weighted one-vs-rest AUC (AUC-OVR), weighted F1-score, and accuracy. Per-class AUC was additionally computed for each of the three healing classes separately using one-vs-rest binarization. All metrics are reported as mean ± standard deviation across folds.

SHAP (SHapley Additive exPlanations) values were computed for TabPFN models fit on the full dataset for classification tasks. A random subsample of up to 200 background observations was used for the SHAP kernel explainer. For classification, SHAP values were computed per class; directional SHAP values (mean signed SHAP per feature per class) were visualized to distinguish features associated with faster versus slower healing. UniProt protein identifiers in feature names were mapped to gene symbols for visualization. The top 25 proteins by maximum absolute SHAP value across any healing class were selected for the summary heatmap. All analyses were implemented in Python using *scikit-learn, xgboost, tabpfn,* and *shap*.

### External validation

To assess whether the proteins identified in the Cox regression analysis showed concordant expression changes at the transcriptomic level, external validation was performed using two independent publicly available RNA-seq datasets retrieved from the NCBI Gene Expression Omnibus (GEO).

The first dataset comprised bulk RNA-seq data from 10 diabetic foot ulcer individuals’ skin and 11 healthy controls skin (GSE199939 (20)). Raw count matrices and associated sample metadata were loaded directly. Samples were assigned to two conditions: DFU and healthy. Genes with fewer than 10 counts in at least 3 samples were excluded prior to analysis. Differential expression analysis was performed using DESeq2, contrasting DFU vs. healthy tissue, with the Benjamini–Hochberg method applied for multiple testing correction (α = 0.05). The second dataset comprised RNA-seq data from DFU ulcer biopsies stratified by healing outcome (GSE134431, (11)), including 6 healed and 7 unhealed DFU patients. Only ulcer tissue samples were retained (perilesional skin samples were excluded). Samples were classified as healer or non-healer. The same low-count pre-filtering criteria were applied (≥10 counts in ≥3 samples). Differential expression analysis was performed using DESeq2 contrasting non-healer vs. healer, with BH correction.

For both datasets, DESeq2 results were filtered to retain only genes whose protein products were identified as significant in the plasma proteomics Cox model (FDR ≤ 0.20). Entrez gene IDs were mapped to gene symbols using the org.Hs.eg.db human annotation database. Results were summarised by log fold change, nominal p-value, and adjusted p-value, and inspected for directional concordance with the proteomic hazard ratio estimates.

### Pseudobulk Differential Expression Analysis

Pseudobulk differential expression was performed on the Theocharidis *et al*. (12) scRNA-seq dataset (GEO accession GSE165816). Cell-level counts were aggregated by patient and cell type. Differential expression between healers and non-healers was performed using DESeq2 with Wald test, applying Benjamini-Hochberg correction. For aggregated analysis, counts were summed across all 21 cell types per patient. For cell-type-resolved analysis, each cell type was analysed separately. Only genes with ≥10 counts in ≥3 patients were retained. Directional concordance with Cox HRs was assessed for the 27 DEPs.

---

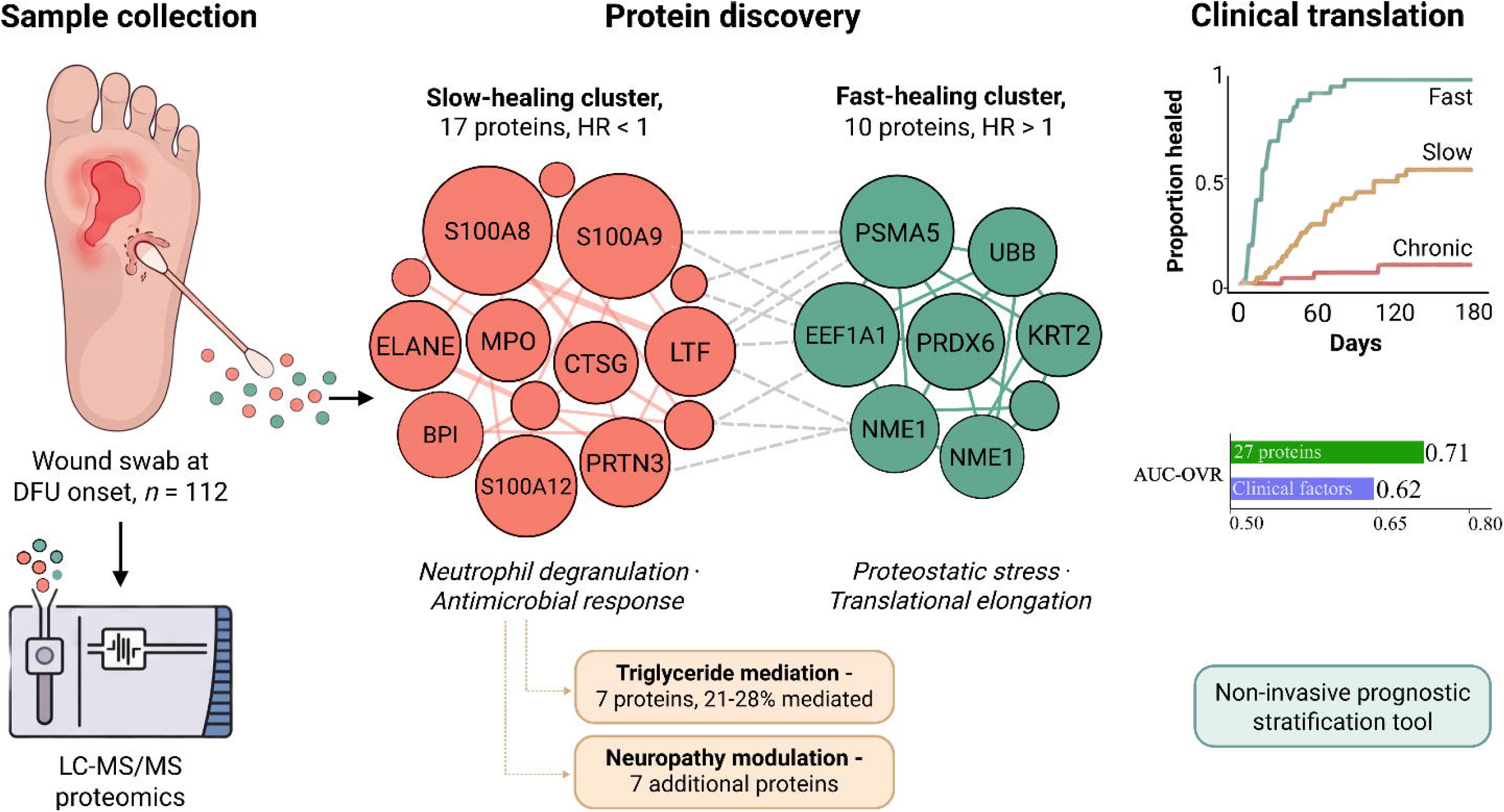

