## Supplementary Results for "Noninvasive Wound Swab Proteomics Enables Prognostic Stratification of Diabetic Foot Ulcer Healing: The HEAL-DFU Proof-of-Concept Study"

**Immune Compositional Shifts Across Healing Groups**

ssGSEA identified a trend toward healing-enriched fibroblast (HE-Fibro) depletion in chronic wounds (ANOVA *p*=0.088; slow-healing vs. chronic *p*=0.084) as the sole finding surviving marker-set filtering (Extended Figure 1A, Supplementary Table S9). BayesDeBulk deconvolution revealed eosinophil enrichment in chronic wounds relative to fast-healing patients (51.3% vs. 30.7%; *p*=0.006) and NK cell depletion in slow-healing vs. fast-healing patients (16.4% vs. 35.5%; *p* =0.002). Dendritic cells were depleted in chronic vs. slow-healing wounds (19.6% vs. 40.6%; *p* =0.008; Extended Figure 2B, Supplementary Table S10). These findings place the elevated SH_C_ neutrophil protein burden within a broader failure of inflammatory resolution: NK cells and dendritic cells contribute to clearance of apoptotic neutrophils and to the transition from the inflammatory to the proliferative phase, and their relative depletion in slow-healing and chronic wounds is consistent with persistence of the neutrophil-derived granule proteins that constitute SH_C_.

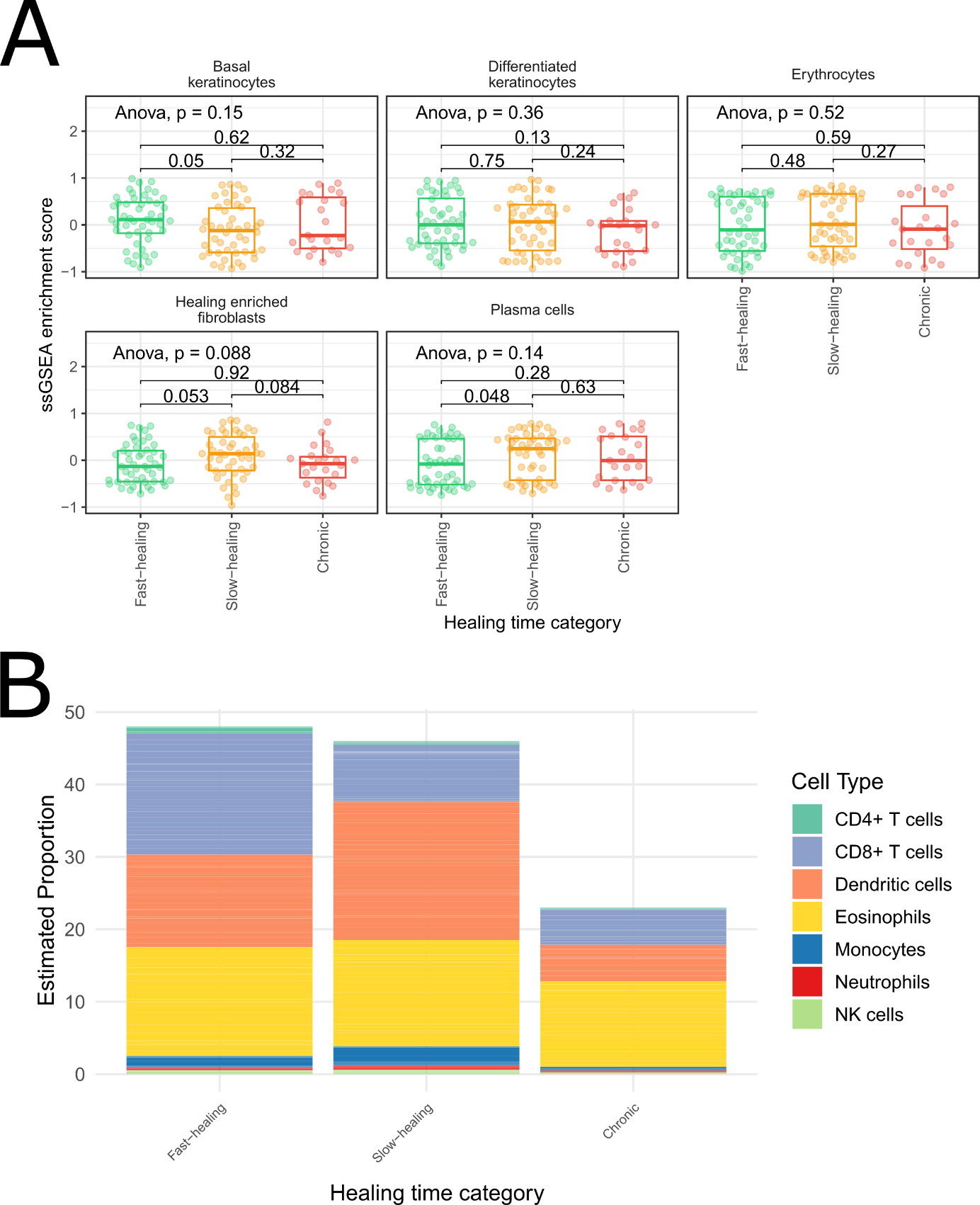

Extended Figure 1: Cell-type deconvolution results across healing time categories. (A) Single-sample gene set enrichment analysis (ssGSEA) enrichment scores for five cell-type signatures: basal keratinocytes, differentiated keratinocytes, erythrocytes, healing-enriched fibroblasts, and plasma cells; stratified by healing time category (Fast-healing, Slow-healing, Chronic). Box-plots show median and interquartile range with individual data points overlaid. Pairwise p-values from post-hoc comparisons and overall ANOVA p-values are annotated. (B) Stacked bar chart showing estimated proportions of immune cell types (CD4+ T cells, CD8+ T cells, dendritic cells, eosinophils, monocytes, neutrophils, NK cells) across the three healing time categories, derived from cell-type deconvolution of the proteomic data.

**External Validation**

We identified 15 out of the 27 DEPs in the protein-protein interaction network provided in Wang *et al* results (1), even do Wang *et al* performed proteomics in a different context (healthy control *vs* DFU) and using a different technology. Specifically, they reported neutrophil degranulation and neutrophil activation as the dominant biological processes in the DFU wound proteome, overlapping with our slow-heal cluster proteins (S100A8/9/12, MPO, CTSG, PRTN3, BPI, CORO1A, VIM, LTF, DEFA3, SERPINB1, TAGLN2). Seemingly Soto *et al* (2) identified a small overlap with our results, particularly those associated with keratinocyte activation markers and extracellular vesicle-associated stress (MPO, PRTN3, CTSG, KRT2).

In GSE199939 (DFU vs. healthy skin; n=21), 11 of 27 proteins were differentially expressed (FDR<0.05, Supplementary Table S11). Three SH_C_ proteins (ELANE, CTSG, and GSN) were significantly lower in DFU tissue than healthy skin, whereas PSMA5 and TAGLN2 (FH_C_) were higher in DFU tissue. Because this dataset contrasts disease state (DFU vs. healthy skin) rather than healing trajectory within DFU, these directions are not directly comparable with our hazard ratios; they do, however, indicate that some signature proteins are dysregulated in the DFU wound environment rather than providing directional replication. In GSE134431 (healer vs. non-healer DFU; n=13), the underpowered design (n=13 total) precluded FDR-level replication; non-significant directional trends were observed for CORO1A, SERPINB1, and VIM.

Aggregated pseudobulk differential expression across all cell types in the Theocharidis *et al*. (2022) scRNA-seq cohort identified 1,208 genes with significant difference between healers and non-healers (FDR<0.05), with 73.4% upregulated in healers, opposite to the SH_C_ proteomic directionality (Supplementary Table S12). Three SH_C_ DEPs were concordantly upregulated in the wound tissue of healers: S100A9 (log2FC=+1.79; padj=1.21×10⁻⁹), S100A8 (+1.44; padj=4.76×10⁻⁶), and PGD (+0.49; padj=9.27×10⁻⁵), and ANXA3 reached FDR-level significance in M1 macrophages (log2FC=+2.52; q=0.039), while EEF1A1 was lower in in the tissue of healers (log2FC=−0.19; padj=3.75×10⁻⁵). Each of these directions is opposite to the corresponding HR, indicating that transcript abundance in viable wound tissue does not mirror the extracellular protein burden measured in wound swabs.
