## Supplementary Figures for "Noninvasive Wound Swab Proteomics Enables Prognostic Stratification of Diabetic Foot Ulcer Healing: The HEAL-DFU Proof-of-Concept Study"

**Supplementary Figure 1**

Gene Ontology Biological Process (GO:BP) overrepresentation analysis results for the 27 differentially expressed proteins, grouped by pathway cluster using enrichment-based and taxonomy-based hierarchical clustering. Two set of bars are included for each functional cluster, one representing the number of individual pathways included in the cluster and another set showing the number of unique relevant proteins represented by each cluster. Bar color encodes the directionality score of the contributing proteins: green indicates Cluster 1 proteins (faster healing, HR > 1) and red indicates Cluster 2 proteins (slower healing, HR < 1); intermediate colors reflect mixed contributions. Bar opacity encodes statistical significance, scaled by the −log₁₀ harmonized mean p-value (HMP) across all enriched pathways within each cluster, as indicated in the right-hand legend.

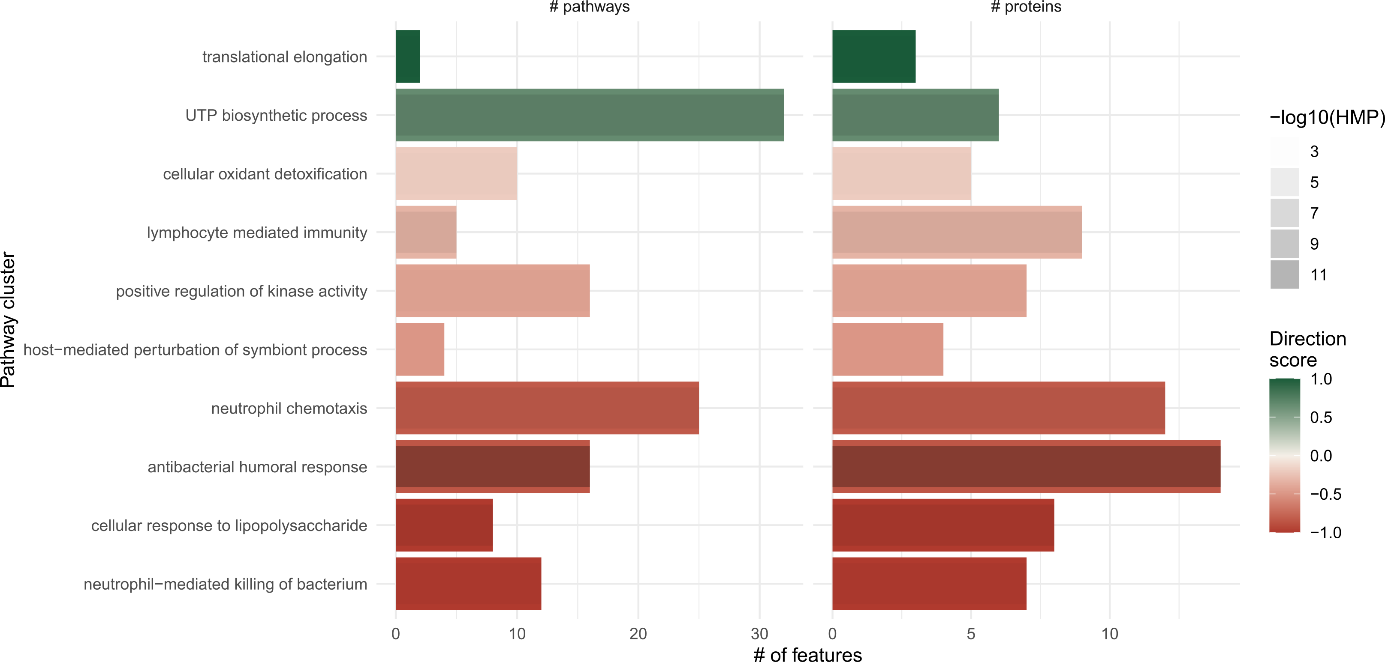

**Supplementary Figure 2**

Functional enrichment network of proteins associated with wound healing outcome obtained from aPEAR pipeline. Each node represents a cluster of functionally related Gene Ontology (GO) biological process terms, with node size proportional to the number of terms in the cluster (cluster size scale: 10–30 terms). Node color indicates the direction score, ranging from +1 (dark green; associated with faster healing) to −1 (dark red; associated with slower healing), with intermediate scores shown in lighter shades. Edges between nodes represent semantic similarity between GO term clusters. Representative GO terms are labelled for each cluster.

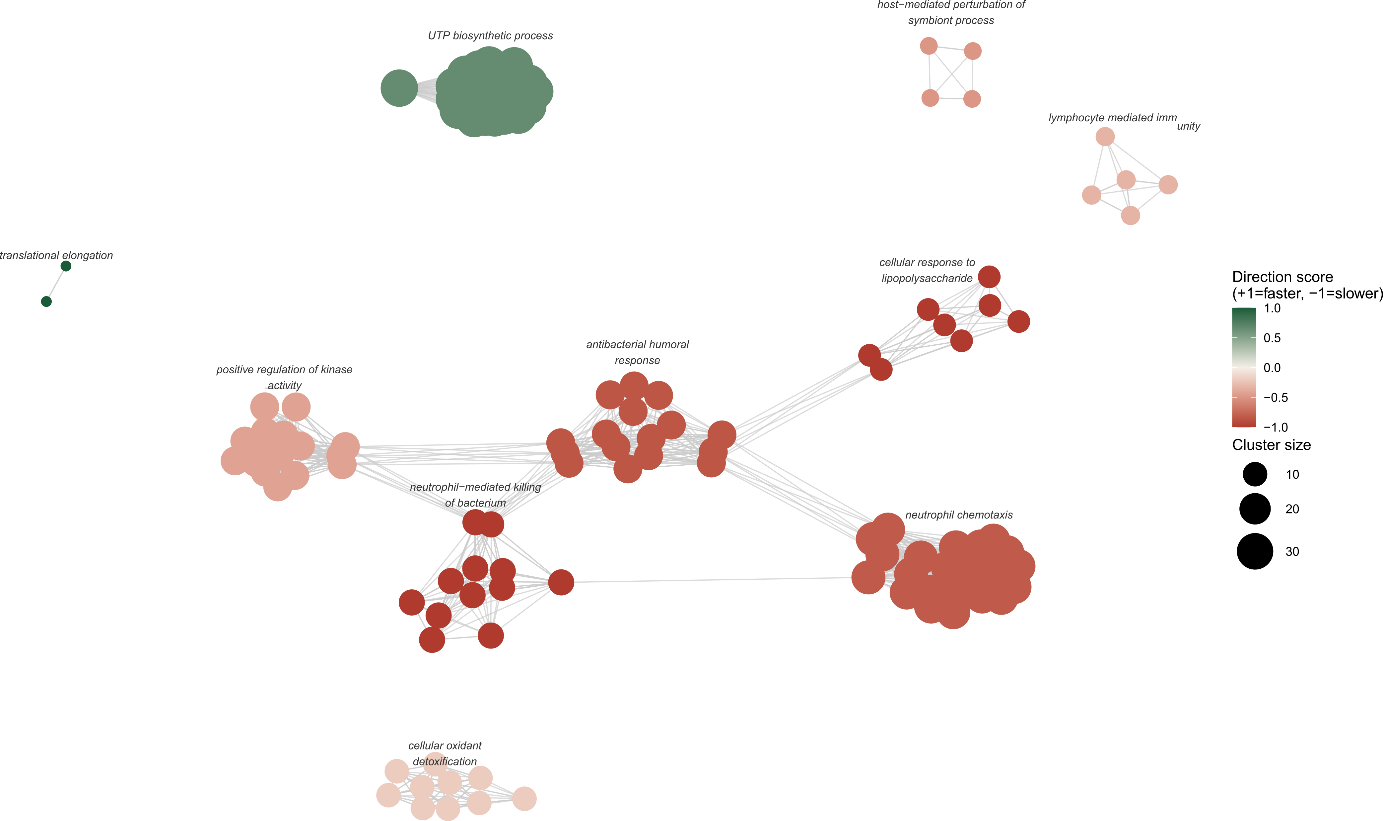

**Supplementary Figure 3**

Forest plot of univariable Cox proportional hazards regression for the 27 signature proteins. Each point represents the hazard ratio (HR) for time-to-healing, with horizontal bars indicating 95% confidence intervals. Proteins are ordered by HR magnitude. Colour indicates FDR group: dark red for 0.05 ≤ FDR < 0.1, lighter red for 0.1 ≤ FDR ≤ 0.2.

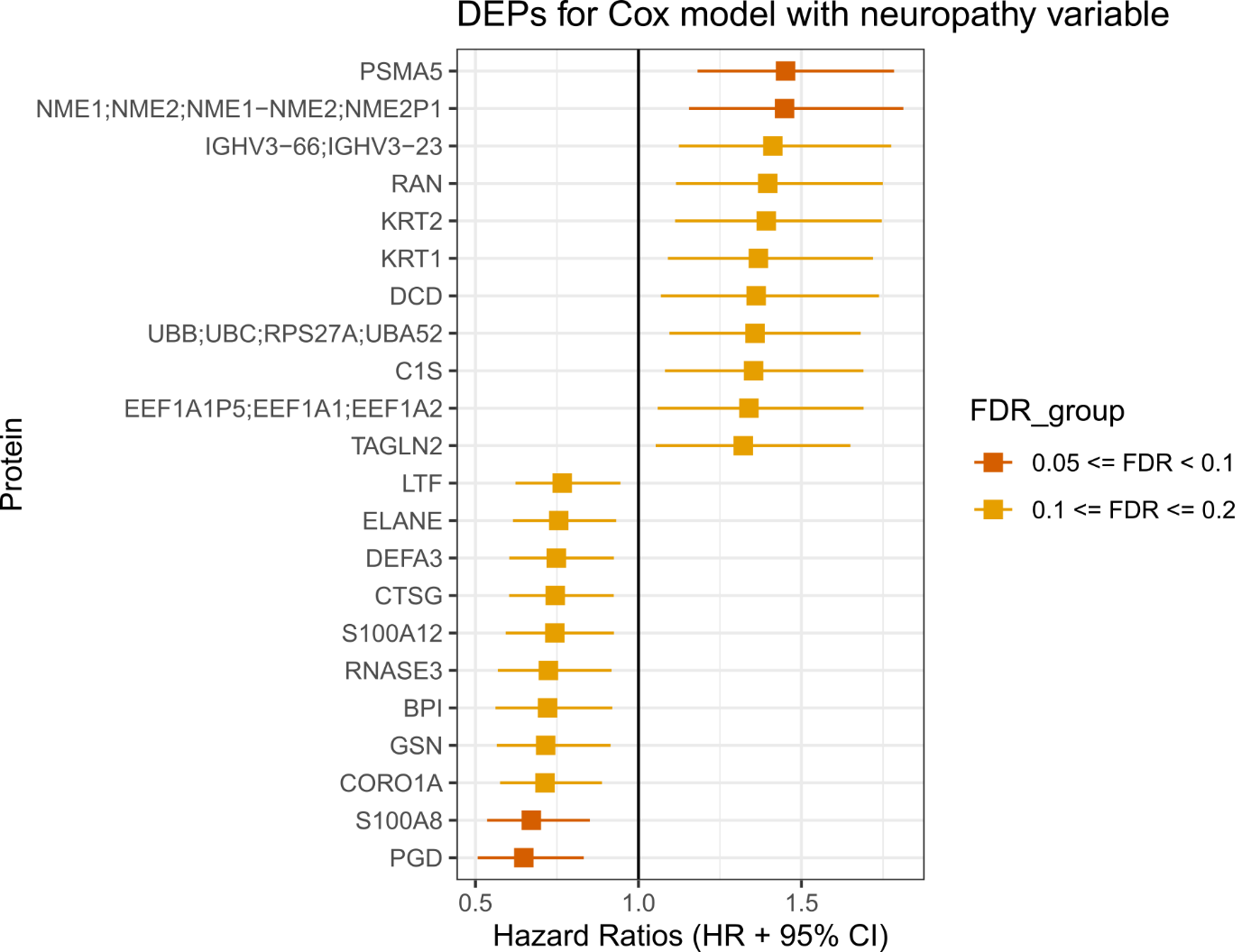
